# Hypoferremia predicts hospitalization and oxygen demand in COVID-19 patients

**DOI:** 10.1101/2020.06.26.20140525

**Authors:** Theresa Hippchen, Sandro Altamura, Martina U. Muckenthaler, Uta Merle

## Abstract

**Background:** Iron metabolism might play a crucial role in cytokine release syndrome in COVID-19 patients. Therefore we assessed iron metabolism markers in COVID-19 patients for their ability to predict disease severity.

**Methods:** COVID-19 patients referred to the Heidelberg University Hospital were retrospectively analyzed. Patients were divided into outpatients (cohort A, n=204), inpatients (cohort B, n=81), and outpatients later admitted to hospital because of health deterioration (cohort C, n=23).

**Results:** Iron metabolism parameters were severely altered in patients of cohort B and C compared to cohort A. In multivariate regression analysis including age, gender, CRP and iron-related parameters only serum iron and ferritin were significantly associated with hospitalization. ROC analysis revealed an AUC for serum iron of 0.894 and an iron concentration <6µmol/l as the best cutoff-point predicting hospitalization with a sensitivity of 94.7% and a specificity of 67.9%. When stratifying inpatients in a low- and high oxygen demand group serum iron levels differed significantly between these two groups and showed a high negative correlation with the inflammatory parameters IL-6, procalcitonin, and CRP. Unexpectedly, serum iron levels poorly correlate with hepcidin.

**Conclusion:** We conclude that measurement of serum iron can help predicting the severity of COVID-19. The differences in serum iron availability observed between the low and high oxygen demand group suggest that disturbed iron metabolism likely plays a causal role in the pathophysiology leading to lung injury.

**KEY POINTS:**

1. Iron metabolism parameters are severely altered in COVID-19 patients.
2. Measurement of serum iron can help predicting the severity of COVID-19.

## INTRODUCTION

Since iron is a critical cofactor for proteins involved in many fundamental biological processes, including DNA/RNA synthesis and ATP generation, viruses, most likely including Coronaviruses, essentially rely on iron to replicate in host cells.^1^ Because both, host and pathogen require iron, the host innate immune response carefully orchestrates iron metabolism to limit iron availability during times of infection. To date little is known about the interaction of the newly discovered coronavirus SARS-CoV-2 with host iron metabolism during Coronavirus disease 2019 (COVID-19).

Patients infected with SARS-CoV-2 may die due to an excessive response of their immune system, hallmarked by an abnormally high release of circulating cytokines, termed cytokine release syndrome (CRS). CRS plays a major role in the deterioration of COVID-19 patients, from pneumonia through acute respiratory distress syndrome (ARDS), cumulating in systemic inflammation and ultimately multi-system organ failure.^2^ This phenomenon of a plethora of cytokines wreaking havoc throughout the body is vividly referred to as “cytokine storm”. Cytokines involved in the “cytokine storm” in COVID-19 patients, include IL-6, IL-1, IL-2, IL-10, and TNF-α. However, IL-6 seems to play the most prominent role, whereby increased levels in the serum correlate with respiratory failure, ARDS, and adverse clinical outcomes.^3^ IL-6 is a pleiotropic cytokine involved in eliciting the acute-phase response in the liver, in B-cell proliferation and antibody production, in T-cell differentiation and cytotoxicity, and in hepcidin synthesis in the liver.^4^ Hepcidin is the master regulator of iron homeostasis. By degrading its target receptor ferroportin, hepcidin controls dietary iron absorption and iron release from iron-recycling macrophages.^5^ During states of infection or inflammation hepcidin levels increase restricting iron availability in the plasma. The resulting hypoferremia is an integral part of the host defense mechanism. Other than IL-6, several other cytokines such as IL-1,^4^ IL-22,^6^ and interferon α,^7^ contribute to increasing hepcidin expression.

In addition to the regulation of iron metabolism via hepcidin, circulating cytokines such as IL-1 and tumor necrosis factor (TNF) increase synthesis of the iron storage protein ferritin.^8,9^ Consequently, more iron is retained predominantly in the reticuloendothelial system that handles most of the iron recycled from damaged red blood cells. The resulting hypoferremia disturbs erythropoiesis and iron uptake in most organs.^10,11^

As the control of iron metabolism is crucial during infections in general and because several cytokines involved in the “cytokine storm” in COVID-19 are strong regulators of iron metabolism, understanding of iron homeostasis in COVID-19 is likely to be a relevant piece in the puzzle of COVID-19 disease. In this study, we noted that severe COVID-19 cases not only show higher levels of inflammatory markers than mild cases, but also that iron metabolism parameters greatly differed depending on disease severity.

Therefore we focused our attention on iron-related parameters in COVID-19 patients and showed that (1) severely ill COVID-19 patients show marked hypoferremia, (2) the level of hypoferremia predicts disease severity (reflected in hospitalization requirement), (3) hypoferremia only partially correlates with hepcidin and inflammatory markers like C-reactive protein (CRP) and IL-6, and (4) immunomodulatory therapeutic agents (e.g. IL-1R blockade, IL-6R blockade and immunoglobulins)^3,12^ show a rapid and strong effect on hypoferremia in COVID-19 patients.

## RESULTS

### Baseline characteristics of patient cohorts

Our outpatient cohort (cohort A) consists of 204 outpatients who remained home throughout the entire course of the disease. 23 patients were initially followed as outpatients but later had to be admitted to the hospital because of clinical worsening. These patients are summarized as cohort C. In addition, we analyzed an inpatient cohort (cohort B) consisting of 81 patients. Table 1 shows the baseline characteristics and treatments of all three cohorts. In cohort B, 48 patients (59.3%) had low oxygen demand (no or low-flow oxygen requirement) while 33 patients (40.7%) showed high oxygen demand (high flow oxygen or invasive ventilation). 14 (17.3%) inpatients of cohort B died, while all patients in cohort A and C survived. Table 2 depicts lab values of patients in all three cohorts. Some of the outpatients of cohort A and C were followed on more than one outpatient visit and data of all time-points are included in laboratory analyses shown in table 2. This explains the results of 415 visits in 204 patients of cohort A and 48 visits in 23 patients of cohort C. The inpatient cohort B differs significantly from Cohort A in all laboratory parameters analyzed.

**Table 1.**
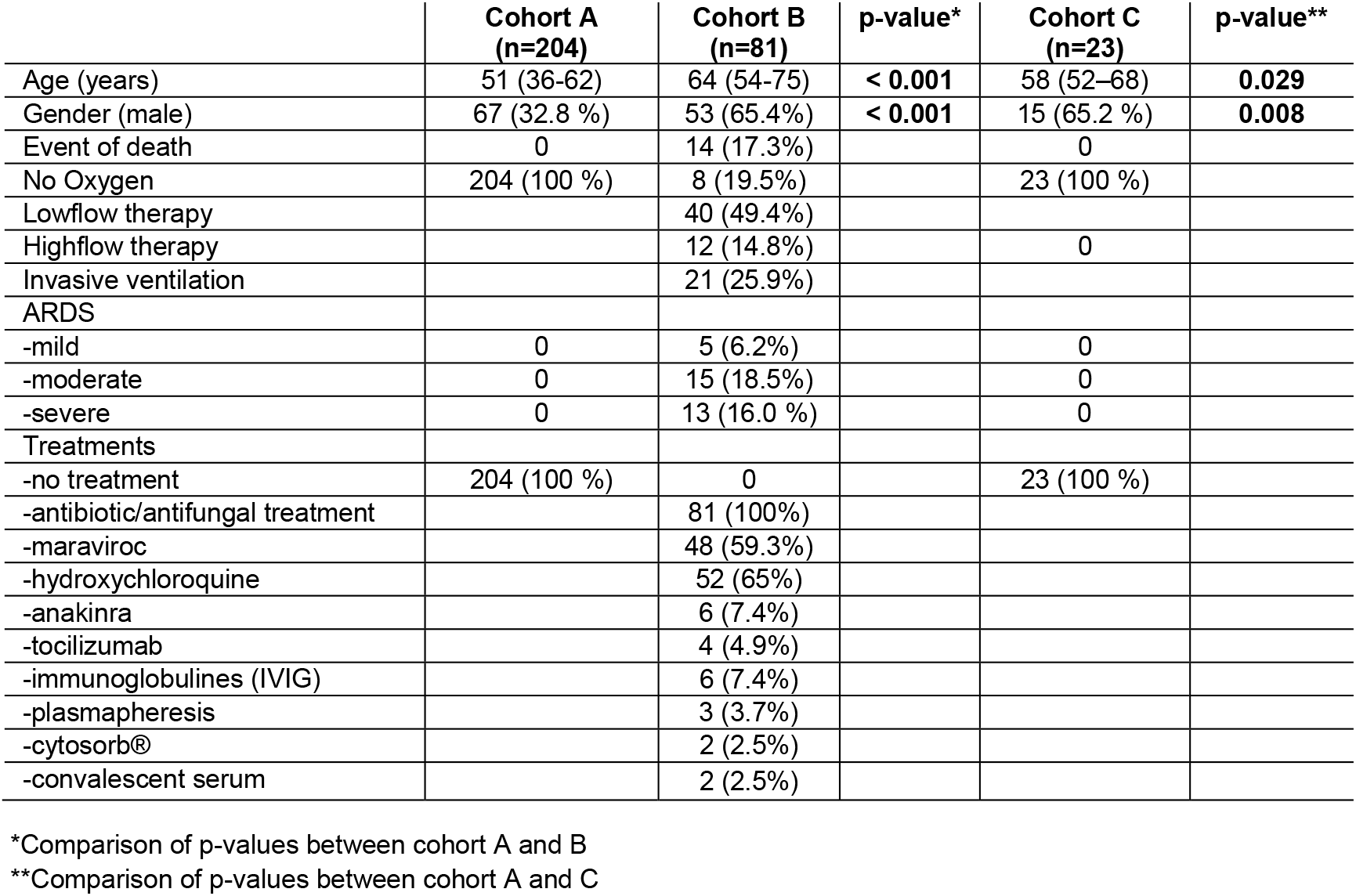
Baseline characteristics of study population. Results are presented as numbers (with %) or median (with interquartile range)

**Table 2.** Laboratory findings of the study cohorts. Results are presented as median (with interquartile range)

|  | **normal range** | **Cohort A (n=204)** | **Cohort B (n=81)** | **p-value\*** | **Cohort C (n=23)** | **p-value\*\*** |
| --- | --- | --- | --- | --- | --- | --- |
| Hemoglobin, g/dL | 13-17 | 14 (13.2-14.9) | 13.3 (11.9-14.7) | **0.003** | 14.3 (13.3-15.4) | **n.s.** |
| White blood cell count, x10<sup>9</sup> per L | 4-10 | 5.8 (4.6-7.3) | 6.4 (4.6-9.0) | **0.033** | 5.3 (3.4-7.3) | **n.s.** |
| Lymphocyte count, x10<sup>9</sup> per L | 1.0-4.8 | 1.5 (1.1-1.9) | 0.8 (0.6-1.1) | **<0.001** | 1.0 (0.7-1.4) | **<0.001** |
| CRP, mg/L | <5 | 13.2 (2.0-40.8) | 89.3 (37.6-148.1) | **<0.001** | 46.4 (21.8-106.8) | **<0.001** |
| Pprocalcitonin, ng/mL | < 0.05 | 0.05 (0.05-0.06) | 0.10 (0.06-0.30) | **<0.001** | 0.06 (0.05-0.11) | **<0.001** |
| Interleukin-6, pg/mL | <15 | n.a | 43.1 (15.8-83.8) |  | n.a. |  |
| Ferritin, µg/L | 30-300 | 227 (83-569) | 777 (341-1339) | **<0.001** | 741 (404-935) | **<0.001** |
| Iron, µmol/L | 14-32 | 8.6 (5.0-14.9) | 2.6 (1.8-3.9) | **<0.001** | 3.2 (2.4-4.6) | **<0.001** |
| Transferrin, g/L | 2.0-3.6 | 1.9 (1.6-2.3) | 1.5 (1.2-1.7) | **<0.001** | 1.7 (1.6-1.9) | **0.012** |
| TfSat, % | 16-45 | 19 (12-28) | 7 (5-11) | **<0.001** | 8 (6-10) | **<0.001** |
| Hepcidin, ng/mL |  |  | 91.4 (59.6-133.7) |  |  |  |
| LDH, U/L | <317 | 286 (232-360) | 384 (308-508) | **<0.001** | 348 (272-374) | **0.005** |
| AST, U/L | <37 | n.a. | 42 (32-77) |  | n.a. |  |
| ALT, U/L | <35 | n.a. | 36 (27-57) |  | n.a. |  |
| Creatinine, mg/dL | 0.6-1.2 | 0.74 (0.65-0.88) | 0.88 (0.69-1.04) | **<0.001** | 0.92 (0.79 – 1.11) | **<0.001** |
| BUN, mg/dL | <45 | 25 (20-31) | 28 (21-44) | **0.001** | 29 (24 – 34) | **0.005** |
| NT-pro-BNP, ng/L | <125 | 99 (54-185) | 247 (109-849) | **<0.001** | 85 (46-154) | **n.s.** |
| Troponin-T, pg/mL | <14 | n.a. | 11.5 (6.0-22.3) |  | n.a. |  |

\* Comparison of p-values between cohort A and B
\*\*Comparison of p-values between cohort A and C
Laboratory values in cohort B were obtained at day of admission. Laboratory values in cohort A and C were obtained during outpatient visits. Cohort A and C: All available values were included in analysis. Cohort B: only values of the day of admission were included in the analysis.
LDH: lactate dehydrogenase; BUN: blood urea nitrogen; AST; alanine aminotransferase; AST: aspartate aminotransferase; CRP: C-reactive protein; PCT: procalcitonin; TfSat: transferrin saturation; n.a. = not analyzed

### Serum iron levels predict hospitalization in COVID-19 patients

COVID-19 patients show strongly elevated inflammatory markers, such as CRP and IL-6, confirming results of previous studies.^13^ In addition, the iron-related parameters analyzed (ferritin, serum iron, TfSat, transferrin and hepcidin) severely deviated from normal in all cohorts. When comparing our cohorts, outpatients of cohort A showed less severe alterations compared to those outpatients that had to be admitted to the hospital because of clinical worsening (cohort C) (table 2). Similarly, iron markers were altered more severely in the inpatient cohort B compared to outpatients in cohort A (table 2). The differences in iron-related parameters in part were gender-specific, with females of cohort A deviating less from the normal range for serum iron, transferrin and ferritin compared to males, while in cohort B and C only ferritin was significantly higher in males (figure 1).

**Figure 1.**
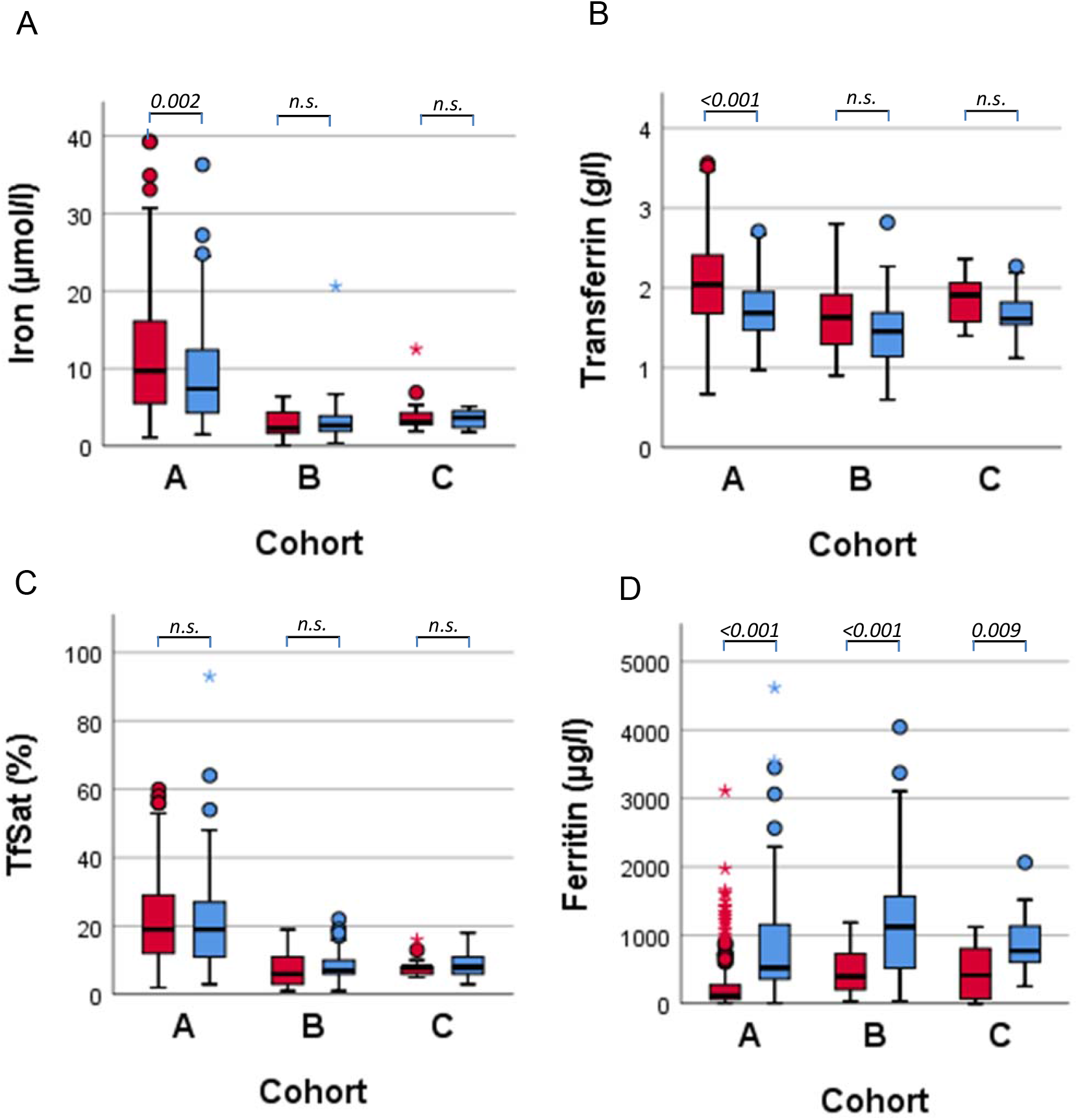
Boxplots representing iron metabolism parameters in female (in red) and male (in blue) patients of cohort A, B and C. Cohort A: 204 outpatients (with 1 to 8 visits per patient, in summary 415 visits); Cohort B: 81 patients with lab values at day of admission; Cohort C: 23 outpatients with clinical worsening and subsequent hospital admission in the course of the disease (with visits up to 9 days before hospital admission and 1 to 6 visits per patient, in summary 48 visits). Figure A: serum iron; B: serum transferrin; C: transferrin saturation; D: serum ferritin. Data are presented as boxplot (median ± interquartile range). Statistical significant distribution of parameters was tested with the Mann-Whitney U test.

Utilizing this unique cohort set up of in- and outpatients, we performed univariate logistic regression analysis for age, gender, CRP, and iron markers with respect to hospitalization requirement of COVID- 19 patients reflecting upon disease severity (table 3). Interestingly, in univariate analysis, all iron metabolism parameters were significantly associated with admission status. In multivariate regression analysis including age, gender, CRP and iron markers only serum iron and ferritin were significantly associated with hospitalization, whereby doubling of serum iron was associated with a 6.7-fold lower odd of hospitalization (adjusted OR per log2 increase in serum iron: 0.15, 95% CI 0.09-0.26, P<0.001) (table 3).

**Table 3.** Univariate and multivariable logistic regression analysis for admission status of COVID-19 patients (cohort A + B)

| Covariate, effect | COVID-19 outpatient (n=204) vs inpatient (n=81) |  |
| --- | --- | --- |
|  | Univariate analysis,<br>OR (95% CI), *P* | Multivariate analysis,<br>aOR (95% CI), *P* |
| Age, $\geq 60$ vs. $<60$ years | 2.26 (1.39-3.70), 0.001 | 0.60 (0.28-1.31), n.s. |
| gender, m vs f | 3.38 (2.05-5.57), $<0.001$ | 1.37 (0.65-2.89), n.s. |
| CRP, per log2 increase* | 2.14 (1.78-2.59), $<0.001$ | 1.20 (0.87-1.66), n.s. |
| Serum ferritin, per log2 increase* | 1.64 (0.14-1.93), $<0.001$ | **1.57 (1.14-2.14), 0.005** |
| Serum iron, per log2 increase* | 0.15 (0.10-0.23), $<0.001$ | **0.15 (0.09-0.26), $<0.001$** |
| transferrin | 0.15 (0.08-0.28), $<0.001$ | 0.90 (0.26-0.32), n.s. |
| Transferrin saturation, per log2 increase* | 0.20 (0.14-0.29), $<0.001$ | |
| Goodness-of-fit test‡ | | $\chi^2=12.97$ (8 df), $P=0.11$ |

*Abbreviations:* COVID-19, Coronavirus-disease 2019; OR, odds ratio; aOR, adjusted odds ratio; CI, confidence interval; df, degrees of freedom.
\*Each one unit increase in log2 corresponds to a doubling in the corresponding parameter.
‡Hosmer-Lemeshow test.

To determine the best discriminating cut-off predicting hospitalization we performed ROC-analysis (figure 2A and 2B). An iron concentration < 6 µmol/l was identified as the best cutoff-point predicting hospitalization. The sensitivity of iron levels at this value was 94.7% with a specificity of 67.9%. The predictive power of iron, expressed as area under the ROC curve (AUC), was 0.894, with a 95% confidence interval of 0.858 – 0.931. The ROC curve AUCs for TfSat, transferrin, ferritin, and CRP were lower compared to iron (0.863 (CI 0.824-0.903), 0.735 (CI 0.676-0.795), 0.725 (CI 0.667-0.784), and 0.838 (CI 0.790-0.886), respectively).

**Figure 2.**
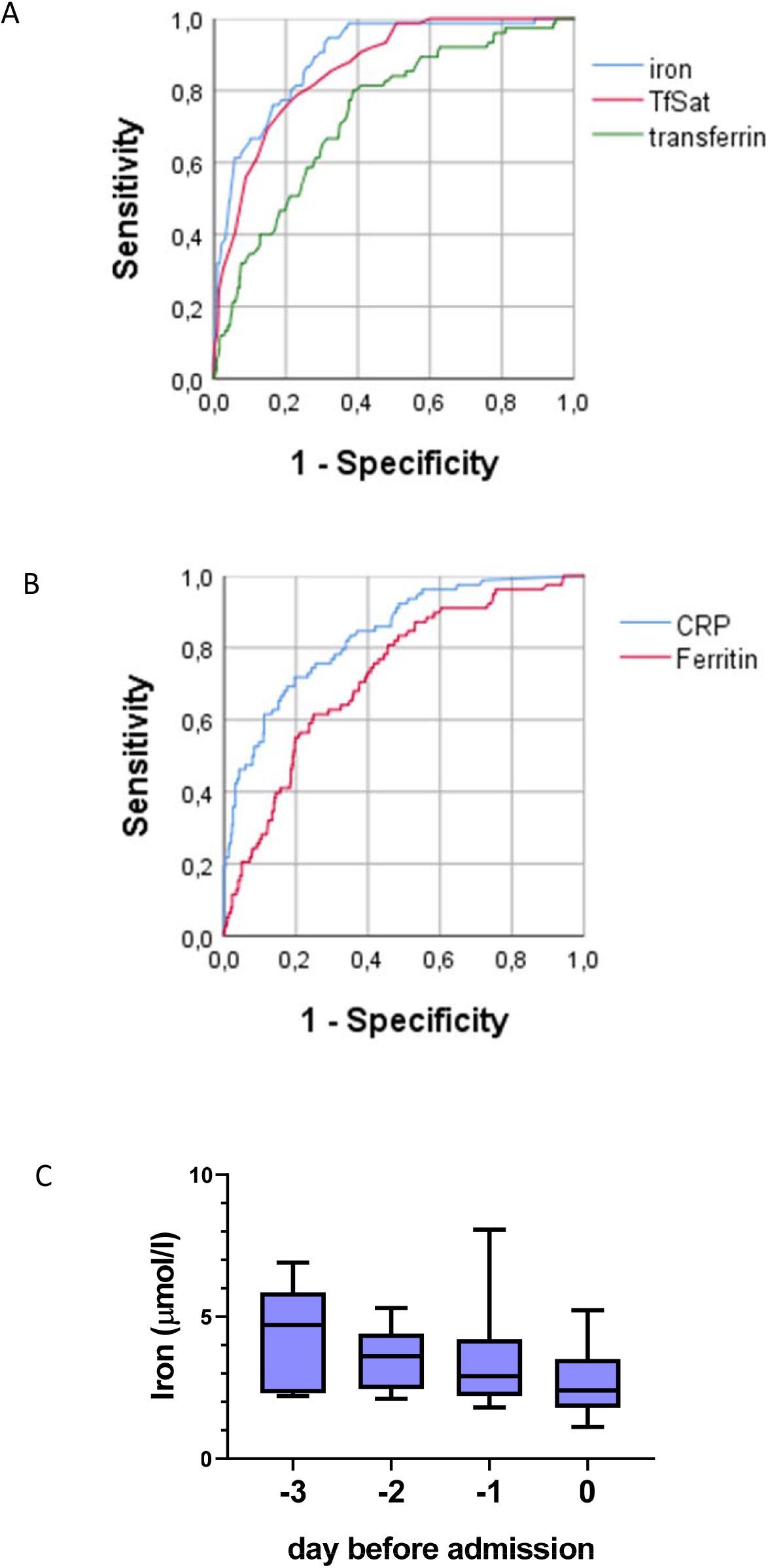
A, ROC curve for cohort A and B for predicting hospitalization for iron, transferrin and TfSat with AUCs of 0.894, 0.735, and 0.863, respectively (and confidence intervals of 0.858-0.931, 0.676-0.795, and 0.824-0.903, respectively). B, ROC curve for cohort A and B for predicting hospitalization for ferritin and CRP with AUCs of 0.725 (CI 0.667-0.784), and 0.838 (CI 0.790-0.886), respectively. C, Course of iron pre-hospitalization for cohort C. Data are presented as boxplot (median ± interquartile range).

Outpatients that were monitored in the days before being admitted to hospital because of increasing disease severity showed a decline of serum iron in the days before admission (figure 2C). However, cohort C (n=23) is small and patients were not monitored at each individual time point; thus, this observation did not reach statistical significance.

Of note, compared to the widely used CRP value, serum iron was predictive in multivariate regression analysis while CRP was not and in ROC analysis the AUC was better for iron than CRP in discriminating in- and outpatients. In conclusion, iron levels are highly predictive for disease progression and hospitalization of COVID19 patients.

### Course of iron-related biomarkers and markers of inflammation during hospitalization

The dynamics of blood parameter alterations may provide interesting insights into COVID-19 disease pathology. Therefore, we focused our analyses on the inpatient cohort and analyzed blood parameters on a daily basis starting from the time of admission (d0) until day 6 (d6). This inpatient cohort was further stratified in a low- (n=48) and a high oxygen demand group (n=33). We investigated whether iron (serum iron, transferrin, hemoglobin) and inflammatory (serum CRP and IL-6) markers differ between patients with low versus high oxygen demand (figure 3). At the day of admission serum iron levels were low in both groups, but increased over the course of the disease in the low-oxygen demand group. By contrast, serum iron levels remained low in the high oxygen demand group. As a result, iron levels differed significantly depending on the oxygen requirement of patients from day three after admission TfSat, an additional indicator of systemic iron availability follows a similar pattern even though the difference does not reach statistical significance at any time point. Severe iron deficiency frequently causes anemia. Consistently, hemoglobin levels are decreased in high oxygen demand patients, suggesting that iron restriction affects the oxygen transport capacity of hemoglobin likely contributing to high oxygen requirement. Additionally, transferrin levels are significantly decreased at the time of hospitalization, which persists over time.

**Figure 3.**
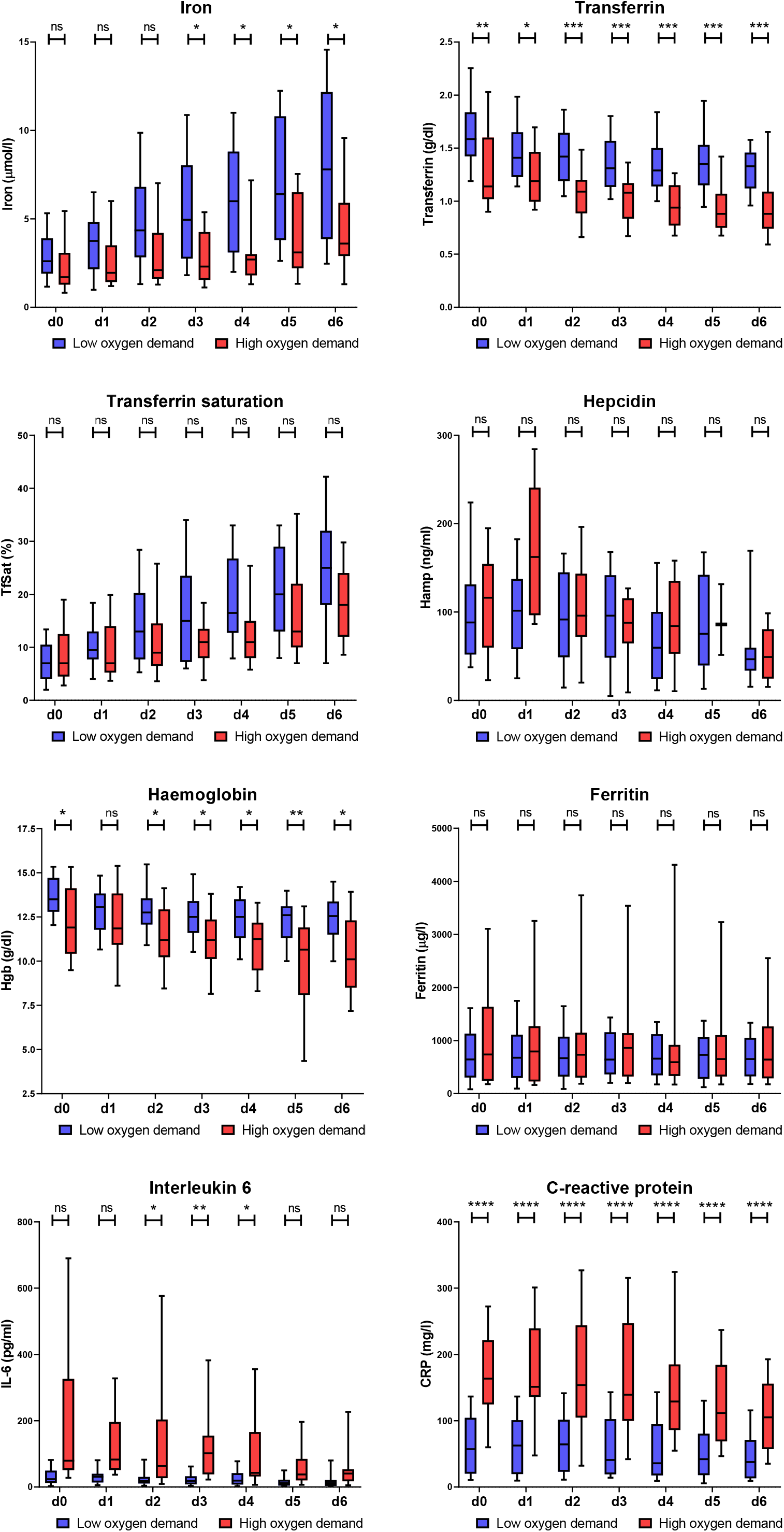
Course of iron and inflammatory parameters in inpatients during the disease course. Serum iron, transferrin, transferrin saturation, hepcidin, hemoglobin (Hgb), ferritin, IL-6 and CRP have been measured at the time of admission (d0) and for 6 following days (d1-d6). Box and whiskers plot represent median with 25^th^ and 75^th^ percentiles (boxes) together with 10^th^ and 90^th^ percentiles (whiskers). Statistical analysis was performed with the Mann-Whitney U test and the p- value has been corrected for multiple comparisons with the Holm method. ns: not significant; *p<0.05; **p<0.01; ***p<0.001; ****p<0.0001

The cytokine IL-6 activates the expression of the peptide hormone hepcidin that causes hypoferremia by blocking iron export from macrophages. Consistently, COVID-19 patients show significantly increased IL-6 and hepcidin levels and a significant correlation of these two parameters. Unexpectedly, hepcidin levels only poorly inversely correlated with serum iron levels. In addition, hepcidin was unable to discriminate between the low and high oxygen demand group, despite the fact that the pro-inflammatory cytokine IL6 and the inflammatory marker CRP are elevated to a higher degree in patients with high oxygen demand compared to those with low oxygen demand. Ferritin levels are highly variable and therefore do not significantly differentiate patients with high and low oxygen demand. Overall, serum iron levels show a high negative correlation with the inflammatory parameters IL-6 and PCT from d0 to d6, while correlation with CRP increases over the time course (figure 4). As expected, iron positively correlates with transferrin saturation and hemoglobin.

**Figure 4.**
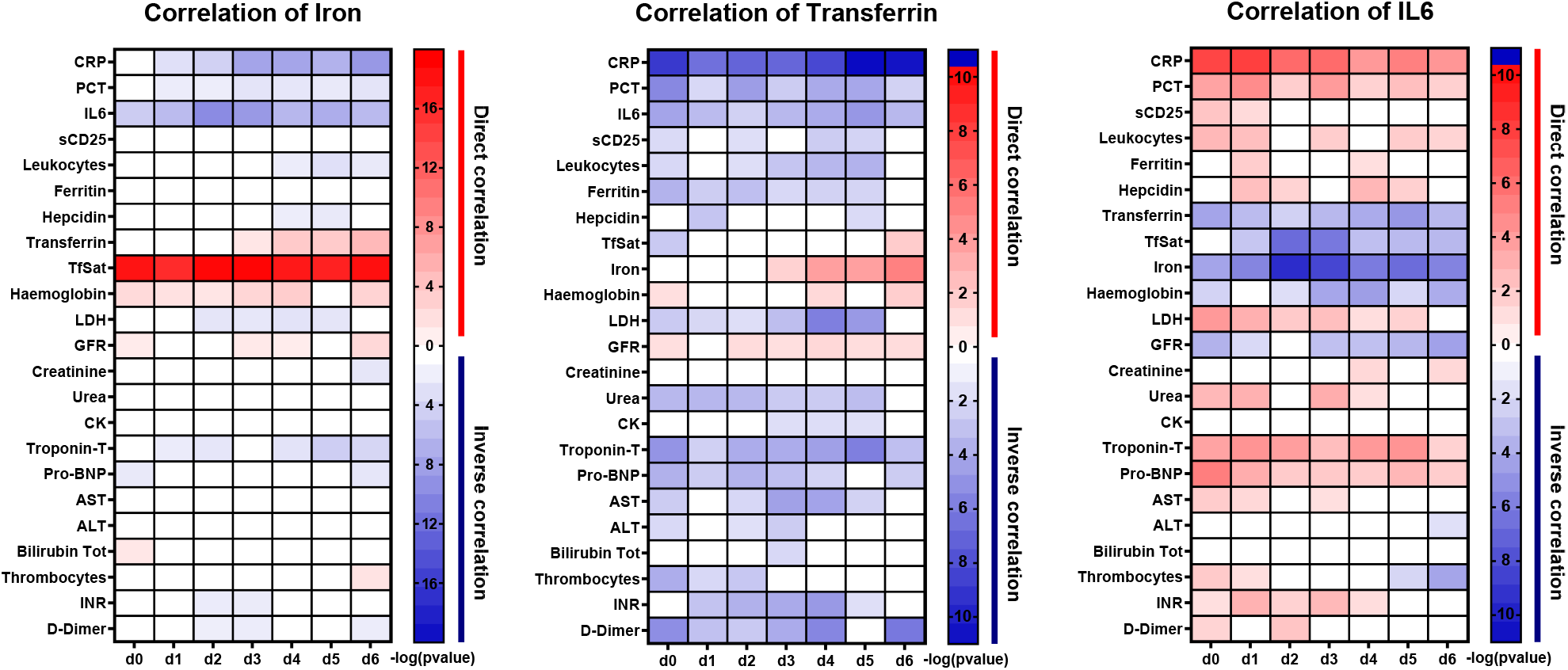
Heatmap of the Spearman’s r correlation analysis of iron, transferrin and IL-6 with the parameters indicated. White boxes indicate a lack of correlation (p>0.05) while in red and blue are reported statistically significant direct and indirect correlations, respectively. The intensity of the color indicates the –log10(pvalue).

We next asked whether hypoferremia correlates with markers for organ damage. Interestingly, low serum iron levels inversely correlate with the cardiac injury marker Troponin-T, while correlations for kidney, liver damage or coagulation were not observed. In addition, transferrin shows a significant inverse correlation with CRP, IL-6, procalcitonin, ferritin, and D-dimers as well as with heart damage parameters Troponin-T and NT-pro-BNP. IL-6 has a similar correlation pattern like transferrin but in addition also negatively correlates with the iron metabolism parameters analyzed.

### Immunomodulatory therapies increase iron availability

Current management of COVID-19 is mainly supportive and approved treatments based on scientific evidence are not available. Main causes of death include ARDS and cytokine storm syndrome therefore therapy with intravenous immunoglobulins (IVIG) or with agents blocking cytokines, like IL-6 receptor antagonist tocilizumab and the IL-1 receptor antagonist anakinra may be efficient.^12,14^ All three treatments have a high anti-inflammatory potential, which allowed us to study the influence of these therapies on iron biomarkers.

In figure 5 the course of iron metabolism parameters before immunomodulatory treatment (defined as day 0) and in the 6 days after treatment is shown. We observed strong effects on serum iron levels and transferrin saturation following therapy with anakinra, tocilizumab and immunoglobulins. When comparing the mean of serum iron levels and transferrin saturation of all patients per treatment group at d1 and d0 these increased 1.8- and 2.1-fold, respectively, after anakinra treatment, 2.0- and 2.2- fold, respectively for tocilizumab treatment, and 2.3- and 2.5-fold, respectively, after immunoglobulin treatment.

**Figure 5.**
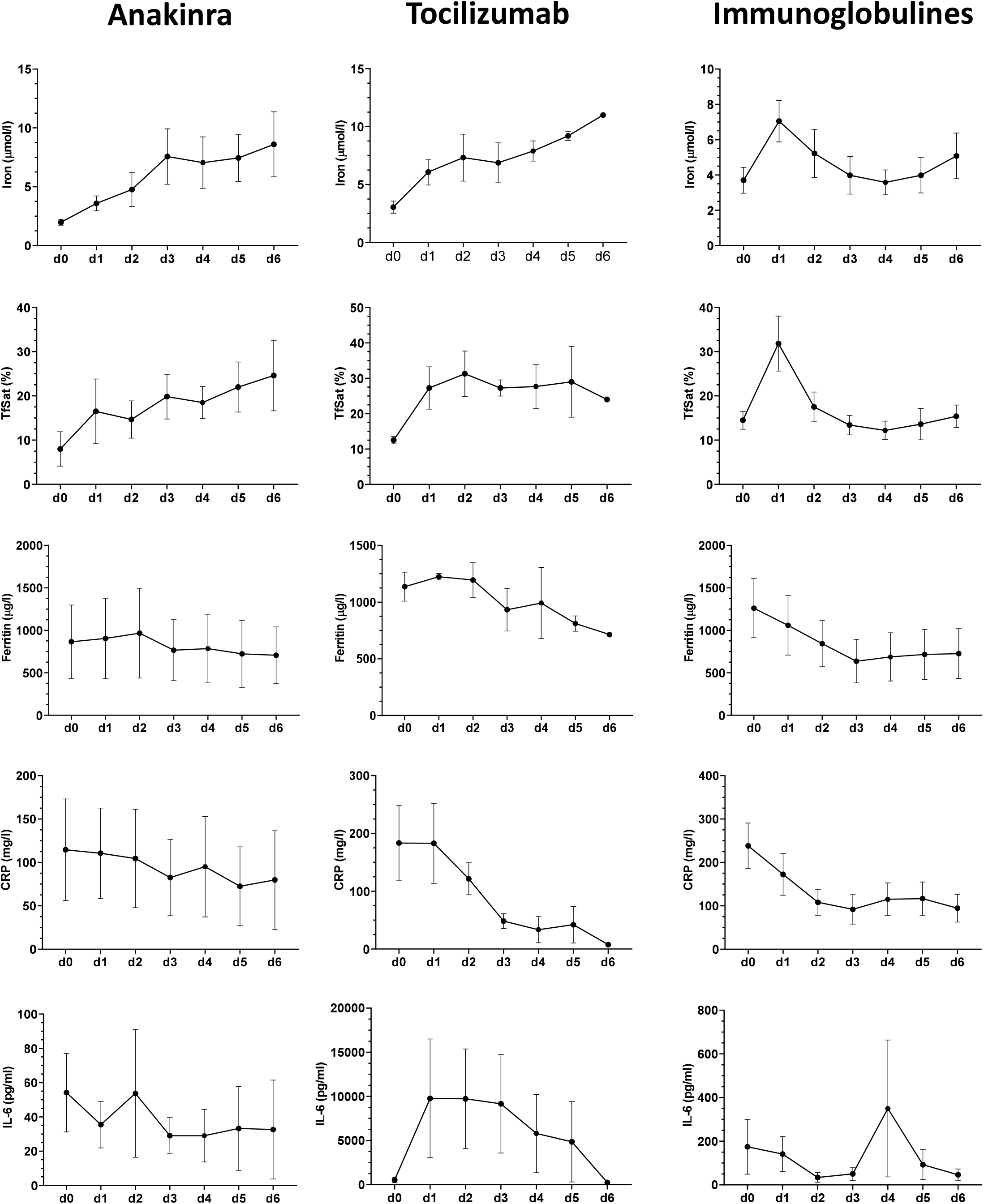
Iron metabolism parameters and CRP before (day0) and in the days after administration of immunomodulatory medication. Displayed are the courses of serum iron, TfSat, ferritin, CRP, and IL-6 for each individual patient under the three different therapies: Anakinra (n=6 patients); Tocilizumab (n=4 patients); intravenous immunoglobulins (IVIG) (n=6 patients). Data are represented as mean±s.e.m.

## DISCUSSION

SARS-CoV-2 infection leads to a broad spectrum of clinical outcomes, spanning from asymptomatic to lethal.^15^ Most of the patients show only moderate symptoms such as weakness, sore throat and fever but in some, the disease progresses to ARDS which in extreme cases can be fatal. Therefore a clinical marker predicting outcome for timely intervention is in high demand. Most studies so far focused on immunologic characteristics as potential markers for monitoring disease severity in COVID-19 patients.^16^

Although iron metabolism plays a central role in the outcome of infections, knowledge about the dynamics of players controlling iron homeostasis in COVID-19 patients is scarce. Our study reveals the predictive value of iron-related markers in patients with confirmed COVID-19 disease. We focus on these parameters as early indicators for disease progression and guidance for therapy decisions in severely affected patients.

We analyzed diagnostic data from our unique patient cohort that is separated into (i) patients with less severe disease taken care of by clinical staff in an outpatient setting (cohort A), (ii) patients initially taken care of in the outpatient setting, but that later deteriorated and had to be admitted to the hospital (cohort C) and (iii) severely ill inpatients (Cohort B). This unique cohort allowed us to determine biomarkers that predict for disease severity reflected by the need for hospitalization and oxygen requirement.

Iron metabolism parameters are disturbed in outpatients and inpatients. Compared to mildly affected patients that remained at an outpatient level, the inpatient cohort shows a significantly more pronounced derangement of iron metabolism parameters. Of note, we observed a gender difference for ferritin in all three cohorts and for serum iron as well as transferrin levels in cohort A, with males being more affected than females. As for COVID-19 a gender difference in disease severity is well recognized, we speculate that lower serum iron levels as well as higher ferritin levels in male patients may be linked with more severe COVID-19 disease in male patients.

In multivariate analysis only serum iron levels and ferritin are significantly predicting disease severity, while CRP, age, gender, as well as transferrin and its saturation are not. Based on a multiple regression analysis model, the odds ratio for hospital admission was 6.7-fold lower per two-fold increase in serum iron. This shows the highly predictive value of serum iron but further hints to a high relevance of altered iron metabolism in the pathogenesis of COVID-19 in the second disease phase typically occurring after the 6th day of disease onset.

Additionally, ROC analysis shows that serum iron levels discriminate outpatients from inpatients, accurately predicting their need for hospitalization. For predicting disease severity reflected in admission status ROC analysis reveals a higher AUC for serum iron (AUC 0.894 with a 95% confidence interval of 0.858 – 0.931) compared to CRP and ferritin levels, that are previously reported COVID-19 severity markers.^17^ An iron concentration < 6 µmol/l was identified as the best cutoff-point predicting hospitalization. The sensitivity of iron levels at this value is 94.7% with an acceptable specificity for a screening marker of 67.9%. In summary, we suggest to assess serum iron parameters in outpatients regularly in order to detect deterioration at an early time point.

The analysis of plasma parameters in 81 patients with COVID-19 hospitalized in the University Hospital Heidelberg provides interesting insights into COVID-19 disease pathology. In this ‘in-patient’ cohort, we analyzed blood parameters on a daily basis starting from the time of admission (d0) until day 6 (d6). Consistent with previous reports, we observed a pronounced ‘inflammatory status’ of COVID-19 patients. Together with inflammation, the most pronounced changes are observed in iron-related parameters. COVID-19 patients show severe hypoferremia, indicated by extremely low serum iron levels and transferrin saturation. Reduced iron availability mildly reduces hemoglobin levels suggesting that this may contribute to the poor oxygenation in severely affected patients. We expect that several mechanisms contribute to the generation of hypoferremia in COVID-19 patients. One major mechanism is the increase of the iron-regulated hormone hepcidin during the acute phase response in the inpatient cohort. In response to inflammation, hepcidin transcription is activated by the cytokine IL-6 via the JAK/STAT signaling pathway. Consistently, during the 6-day time course hepcidin and IL-6 correlate, whereby both initially increase and subsequently decline. Hepcidin triggers degradation of the iron exporter ferroportin thus blocking iron release to the blood stream. Therefore, high amounts of iron derived from the recycling process of senescent red blood cells are retained in macrophages. Both macrophage iron accumulation and inflammation will contribute to the rise in levels of the iron-storage protein ferritin, a marker consistently observed in our in-patient cohort. Iron retention in macrophages due to high hepcidin levels may at least in part explain the severity of hypoferremia in COVID-19 patients. Unexpectedly, Spearman’s correlation analysis, which confirms strong direct correlations between serum iron levels and transferrin saturation and inverse correlations between hepcidin and the inflammatory markers IL-6, PCT and CRP (figure 4), failed to reveal a correlation between serum iron and hepcidin levels. We suspect that the transcriptional repression of the iron exporter ferroportin via pro-inflammatory cytokines may be a driving force causing hypoferremia in COVID-19 patients.^18,19^ In addition, the elevation of hepcidin levels in COVID-19 patients may be inadequate in the context of the extreme inflammation. It is possible that the hypoxic state in COVID-19 patients triggers erythropoietin-dependent expression of the blood hormone erythroferrone in erythroid precursor cells, which in turn represses hepcidin in hepatocytes explaining the poor correlation between hepcidin levels and the degree of hypoferremia in these patients. Thus, the cytokine storm reported in COVID-19 patients may cause hypoferremia by at least three mechanisms, hepcidin-mediated ferroportin degradation, transcriptional downregulation of ferroportin, and elevated iron retention due to high ferritin levels. Of note, COVID-19 patients also show strongly decreased transferrin levels. While literature links inflammation with decreased serum transferrin,^20,21^ the molecular mechanism and biological meaning of this phenomenon are not clear.

As outlined above, high ferritin levels may in part be explained by iron retention in macrophages. We have previously shown that macrophages exposed to heme and/or iron released during hemolysis undergo a phenotypic change that is hallmarked by a pro-inflammatory state.^22^ Heme release from red blood cells is frequently observed in conditions of sepsis. COVID-19 patients show elevated LDH levels, a marker for hemolysis. We therefore propose that COVID-19 patients with severe hypoferremia likely display a high degree of iron accumulation in macrophages that, together with hemolysis may contribute to the severe inflammation observed in these patients.

Another important finding of this study is that iron levels negatively correlate with the myocardial damage marker Troponin-T, suggesting that reduced iron availability could be a co-factor for cardiac stress. The heart is particularly rich in mitochondria that rely on a large amount of iron to produce energy via oxidative phosphorylation. Consistently, iron deficiency has been widely reported as a possible comorbidity for cardiac injury.^23,24^

Similar to hypoferremia, transferrin and IL6 display a strong correlation with inflammatory markers (CRP, PCR and Leukocyte number), markers of tissue-damage (LDH) and cardiac injury (Troponin-T and NT-pro-BNP). However, in addition, these two markers correlate with the glomerular filtration rate and serum urea levels, even though these parameters do not reach pathological level (table 3).

Under low oxygen conditions, erythropoietin stimulates erythropoiesis, a process that requires high amounts of iron.^5^ We next questioned whether the immense reduction in systemic iron availability in COVID-19 patients may be involved in the worsening of ARDS. Using a retrospective approach, we divided cohort B into two subgroups, those with high and those with low oxygen demand. We observed that serum iron levels are lower in patients with high oxygen demand already at the time of admission to the hospital, a difference that increases during the time course and becomes significant three days later. A similar tendency is observed for transferrin saturation at most time points analyzed. Taken together, our data suggest that reduced iron availability is associated with and might even contribute to the progression of ARDS in COVID-19 patients. Additional studies are required to address how iron availability contributes to ARDS.

As hypoferremia is correlated with CRP- and IL-6-levels we wanted to understand the influence of immunomodulatory therapies on iron levels. Of interest, all three analyzed anti-inflammatory treatments showed a rapid and profound influence on serum iron levels already one day after therapy. The fact that serum iron levels increased after all three anti-inflammatory treatments is in line with our hypothesis that inflammation is the major driver of hypoferremia. Interestingly, the influence of anakinra and tocilizumab on ferritin levels was only mild. This may be explained by immune dysregulation in severe COVID-19 patients that has previously been shown to be only partially normalized by tocilizumab.^25^ The fact that in our studies hypoferremia reacts differently than CRP- and IL-6 levels shows that immune dysregulation in COVID-19 is complex and has to be further analyzed in future studies.

In summary we show that measurement of serum iron levels can help in predicting the severity of COVID-19 disease. We propose that both, hypoferremia as well as the expected iron accumulation in macrophages may play a causal role in the pathophysiology of this disease leading to lung injury.

## PATIENTS AND METHODS

### Patient selection

All patients with laboratory-confirmed SARS-CoV-2 infection with an age of 18 years or older that were followed as an out- or inpatient at the University hospital of Heidelberg between March 1st and April 23rd, 2020 were considered for inclusion. Data analysis was approved (number S-148/2020) by the Ethics Committee of the Medical Faculty Heidelberg.

Diagnosis of SARS-CoV-2 infection was based on a positive reverse-transcriptase quantitative polymerase chain reaction (RT-qPCR) to detect the viral genome from individual throat swabs or airway surface liquid.

The outpatient cohort consisted of patients who were visited at regular intervals by medically trained staff at their homes, because they reported relevant symptoms like dyspnea or continuously high fever. The outpatients could either stay at home for the entire course of the disease (cohort A) or were admitted to our hospital because of relevant deterioration (cohort C).

All patients that were admitted to the Internal Medicine department of the University hospital Heidelberg due to SARS-CoV-2 infection were considered for inclusion into the in-patient cohort (cohort B). Additional inclusion criteria for the inpatient cohort were severe symptoms (like severe dyspnea or neurological symptoms) and/or a severe or critical course of COVID-19 at the admission time point. Severe COVID-19 was defined as patients showing one or more of the following characteristics at admission to hospital: respiratory rate ≥30/min, blood oxygen saturation ≤93%, ratio of partial pressure of oxygen in arterial blood over the fraction of inspired oxygen <300mmHg. Patients that had been primarily treated in other hospitals and were secondarily transferred after an external treatment period of >48h were not included in the study cohort. In addition, patients that were only hospitalized for quarantine or psychological reasons were also excluded from the analysis. Based on these criteria 81 inpatients were included for further analysis and 12 patients were excluded.

For all patients’ baseline characteristics, treatments, oxygen requirement, laboratory values, and outcome measures were obtained. Inpatients were stratified in a low and high oxygen demand group. All patients requiring High-Flow-Nasal-Oxygen (HFNO) or invasive ventilation in order to reach a blood oxygen saturation >93% were stratified in the high-oxygen demand group, and all remaining patient in the low-oxygen demand group. Furthermore, hepcidin levels were measured using a commercially available ELISA assay according to the manufacturer’s instructions (DRG Diagnostics).

To study the effects of certain highly immunomodulatory therapeutic agents on iron metabolism we analyzed defined cohorts of patients before and after treatment with anakinra, tocilizumab, and intravenous immunoglobulins (IVIG).

The study was approved by the local ethics committee in Heidelberg (ethics approval number S- 148/2020) and was conducted in accordance with the Declaration of Helsinki.

### Statistical analysis

Data in tables are presented as median with interquartile range (IQR) and as numbers (with percentages) in case of categorical data. For comparison between variables, the Mann-Whitney U-test or Wilcoxon’s test, chi-squared or Fisher’s exact test were used as appropriate. In case of multiple comparisons, the p-value has been corrected with the Holm method. Spearman’s correlation was used to determine correlation of laboratory values. Since CRP levels and iron parameters showed a left-skewed distribution, data was log2 transformed for further analysis. A univariate logistic regression analysis was chosen as the screening method to assess the relationships between putative predictive parameters and the severity of COVID-19, that was defined either by the status of the patients (in- versus outpatient) and for the inpatients by the oxygen demand (low versus high oxygen demand). The predictor variables identified as significant by the univariate logistic analysis were then entered into a multiple logistic regression model to establish which of them could most accurately predict COVID-19 severity including age, gender, and CRP as confounders. Model calibration was evaluated by using the Hosmer-Lemeshow goodness-of-fit test.^26^ The odds ratios (ORs) and their 95% confidence intervals (CIs) for each of the variables were then generated to clarify the respective association of each of the risk factors with the COVID-19 severity. In addition, receiver operating characteristic (ROC) curve analysis with calculation of the area under the curve (AUC) was performed. Data collection and analyses were performed using SPSS 21 (IBM Corp. Armonk, NY, USA) or with GraphPad prism 8 (GraphPad Software, San Diego, CA, USA). For all tests, a p-value < 0.05 was considered to be statistically significant.

## Data Availability

All data will be available upon request after final publication in a peer reviewed journal.

## AUTHORSHIP CONTRIBUTIONS

The rationale was drafted by UM. Patients were treated by UM and TH. Data acquisition was done by TH and UM. Data analyses, interpretation, and figure design were carried out by SA, UM, MUM, and TH. The manuscript was written by UM, MUM, SA, and TH. All authors revised and approved the manuscript.

## Notes

**CONFLIFT OF INTEREST DISCLOSURES** There are no potential conflicts of interest.

**FUNDING** MUM acknowledges funding from the Deutsche Forschungsgemeinschaft (SFB1036, SFB1118) and from the Federal Ministry of Education and Research (NephrESA project Nr 031L0191C).

### Competing Interest Statement

The authors have declared no competing interest.

### Funding Statement

MUM acknowledges funding from the Deutsche Forschungsgemeinschaft (SFB1036, SFB1118) and from the Federal Ministry of Education and Research (NephrESA project Nr 031L0191C).

### Author Declarations

Data analysis was approved (number S-148/2020) by the Ethics Committee of the Medical Faculty Heidelberg.

